# Uptake and determinants of malaria vaccination among under-five children in the Tiko Health District: a community based cross-sectional study

**DOI:** 10.1101/2025.05.08.25327274

**Authors:** Idang Maureen Abiache, Divine Nsobinenyui, Chrisantus Eweh Ukah, Yunika Larissa Kumenyuy, Ngu Claudia Ngeha, Randolf Wefuan, Syveline Zuh Dang, Ndip Esther Ndip, Mirabelle Pandong Feguem, Dickson S. Nsagha

## Abstract

**Background:** Malaria remains a major public health challenge in Cameroon, with children under five years of age being the most vulnerable group. In 2021, the World Health Organization (WHO) recommended the use of the RTS,S/AS01 malaria vaccine in areas of moderate to high malaria transmission. This study assessed caregiver practices and determinants influencing malaria vaccine uptake among under-five children in the Tiko Health District.

**Methods:** A community-based cross-sectional study was conducted from the 1^st^ of March 2025 to the 11^th^ of April 2025 among 410 caregivers of children aged 0–5 years, selected using multistage sampling. Data were collected using a structured questionnaire covering sociodemographic characteristics, vaccine-related practices, and factors influencing uptake. Descriptive statistics were computed, and logistic regression analyses were used to determine factors independently associated with malaria vaccine uptake.

**Results:** Only 32.2% (n = 132) of children had received the malaria vaccine. Among those vaccinated, 72.0% completed all recommended doses, and 82.6% of caregivers reported keeping vaccination records. However, only 24.1% of all respondents demonstrated good overall practices toward the vaccine. Factors associated with uptake of malaria vaccine were, female caregivers (aOR: 4.16, 95% CI: 1.47–11.75), caregivers in health professions (aOR: 2.87, 95% CI: 1.35–5.69), biological parents (aOR: 11.44, 95% CI: 1.52–86.11), and those with monthly income of XAF 51,000–100,000 (aOR: 2.76, 95% CI: 1.68–4.55) had significantly higher odds of vaccination. Trust in health workers (aOR: 6.12, 95% CI: 2.97–12.61) and information from healthcare providers (aOR: 7.60, 95% CI: 3.82–15.08) were also strong predictors and children who had previously suffered from malaria were less likely to be vaccinated (aOR: 0.31, 95% CI: 0.18–0.54).

**Conclusion:** Malaria vaccine uptake in the Tiko Health District remains low, and caregiver practices are generally poor. Factors such as caregiver sex, profession, household income, and access to information and healthcare services significantly influenced uptake. Strengthening caregiver education, improving access to health services, and building trust in healthcare providers are essential for increasing malaria vaccine coverage.

## Background

Malaria continues to be a public health concern in sub-Saharan Africa, particularly affecting children under five years of age. According to the World Health Organization (2023) [1], there were approximately 249 million malaria cases and 608,000 malaria-related deaths globally in 2022, with over 80% of deaths occurring in sub-Saharan Africa. Young children account for more than 70% of all malaria-related fatalities (WHO, 2023) [1]. In Cameroon, malaria is a leading cause of morbidity and mortality among young children. The country reports over seven million malaria cases annually, with children under five being the most affected group [2]. The introduction of the RTS,S/AS01 malaria vaccine, known as Mosquirix, represents a significant advancement in malaria prevention. In October 2021, the WHO recommended the widespread use of this vaccine for children in regions with moderate to high transmission of *Plasmodium falciparum* (WHO, 2021) [3]. Studies from pilot countries conducted in Ghana, Kenya, and Malawi have shown a 30% reduction in severe malaria cases and hospitalizations among children who completed the four-dose series [4]. Cameroon became one of the first countries outside the pilot programs to integrate the malaria vaccine into its routine immunization plan, launching the program in January 2024 [5]. Despite these advances, the success of the malaria vaccine program depends on achieving high coverage and acceptance within communities. Socioeconomic status, cultural norms, and exposure to misinformation also influence decision-making regarding malaria vaccination [6]. Vaccine hesitancy is also a significant factor contributing to the low vaccination coverage observed in many African countries [7]. Furthermore, lack of awareness, financial challenges, cultural beliefs and logistical challenges pose a problem to malaria prevention [8]. Understanding caregiver practices and the factors influencing vaccine uptake is important for the success of the vaccination program. Studies have shown that caregiver education level, attendance at antenatal clinics, and closeness to vaccination centers significantly affect vaccine [9]. In Cameroon, the Ministry of Public Health, in collaboration with global partners are working on malaria vaccine program as part of its integrated malaria control strategy. Parents and caregivers are very important in making decisions about their children’s healthcare, particularly in ensuring vaccination uptake and following recommended immunization schedules [10,11]. Caregiver practices, awareness of vaccine availability, and health-seeking behaviors, are important to ensuring complete and timely vaccination. Therefore, this study, aims to assess the uptake of the malaria vaccine among under-five children in the Tiko Health District. Specifically, it seeks to assess caregivers’ practices toward malaria vaccination and identify factors influencing vaccine uptake.

## Methodology

### Study Design

This was a community-based cross-sectional design aimed at assessing the uptake of the malaria vaccine and its determinants among children aged 0–5 years in the Tiko Health District of the South West Region of Cameroon.

### Study Area

The study was conducted in Tiko Health District (THD), located in Fako Division, South West Region of Cameroon, from the 1^st^ of March 2025 to the 11^th^ of April 2025. Tiko is comprised of eight health areas and serves an estimated population of 55,914 people with diverse ethnic and socioeconomic backgrounds. The health district is characterized by a combination of urban and peri-urban settlements, and it presents unique public health challenges such as poor sanitation and limited access to health services.

### Study Population

The target population consisted of caregivers of children aged 0–5 years residing in the Tiko Health District during the study period.

### Sample Size Determination

The sample size was calculated using the formula for estimating a population proportion with a 95% confidence level [12]:

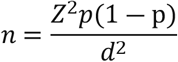

Where;

Z= 1.96 (for 95% confidence level)

P= 0.5 (estimated proportion),

E= 0.05 (margin of error)

This gave us a sample size of 384, which was increased to 410 to account for non-response rate.

### Sampling Technique

A multi-stage sampling technique was used:

Simple random sampling was used to select health areas from within the Tiko Health District.

Probability proportional to size (PPS) was applied to allocate the sample size across the selected health areas.

Systematic sampling was used to select eligible households within each health area. Community health workers assisted in identifying households with children aged 0–5 years.

### Inclusion and Exclusion criteria

Caregivers of under-five children who gave their consent were included in the study while those who were severely ill or had cognitive impairment were excluded.

### Data Collection Tools and Procedures

A structured, pre-tested questionnaire was used to collect data. The questionnaire was divided into four sections: Section A: Socio-demographic information, Section B: Knowledge about the malaria vaccine, Section C: Attitudes towards the malaria vaccine and Section D: Use and practices regarding malaria vaccination

Pre-testing of the questionnaire was conducted in the Mutengene Health Area to ensure clarity, consistency, and reliability. Necessary revisions were made before the full rollout.

Data collection was conducted by the principal investigator and four trained research assistants. For literate participants, questionnaires were self-administered, while illiterate participants were interviewed by research assistants who read questions aloud and recorded responses.

### Data Management and Analysis

Data from completed questionnaires were checked for completeness and accuracy, coded, and entered into SPSS version 26 for analysis. Descriptive statistics were used to summarize data: means and standard deviations for continuous variables, and frequencies and percentages for categorical variables. Practices scores were computed by assigning 1 point for each correct or appropriate response and 0 for incorrect or inappropriate ones. A 60% cutoff was used to categorize respondents into “good” or “poor” groups [13]. Bivariable analysis (using simple logistic regression) was conducted to assess associations between independent variables and vaccine uptake. Multivariable analysis was done using multiple logistic regression, performed to identify independent predictors of malaria vaccine uptake. Statistical significance was set at p < 0.05.

### Ethical Considerations

Ethical clearance was obtained from the Southwest Ethics Committee for Human Health Research in Buea (135/CRERSH/SW/C/02/2025). Administrative authorization was secured from the Faculty of Health Sciences of the University of Douala, and the Tiko District Health Service. Participants were briefed on the study’s purpose, benefits, and voluntary nature. Written informed consent was obtained from all participants. Confidentiality and anonymity were strictly maintained by using unique codes instead of names on questionnaires.

## Results

### Socio-demographic characteristics

The mean age of the 410 caregivers was 33.6 and the standard deviation was 8.9. A total of 219 (53.4%) of caregivers were within the age group 21-35 years and 341 (83.2%) of them were females. Secondary education was the dominant educational level 160 (39.0%) and 340 (82.9%) were Christians. A majority 249 (60.7%) were married and 158 (38.5%) were self-employed. A vast majority 329 (80.2%) of the caregivers were direct parents of the under children and 391 (95.4%) were non-smokers (Table 1)

**Table 1:**
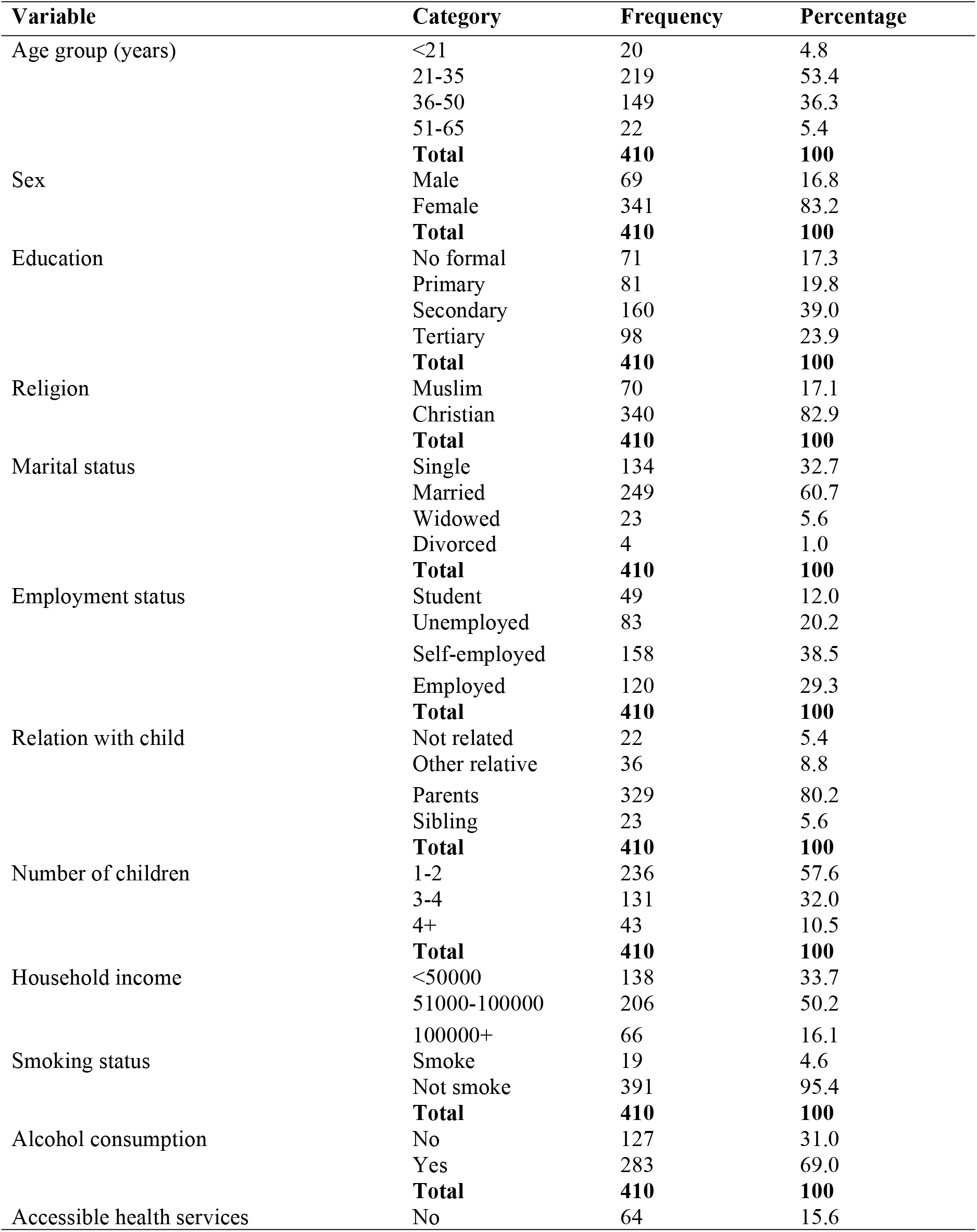
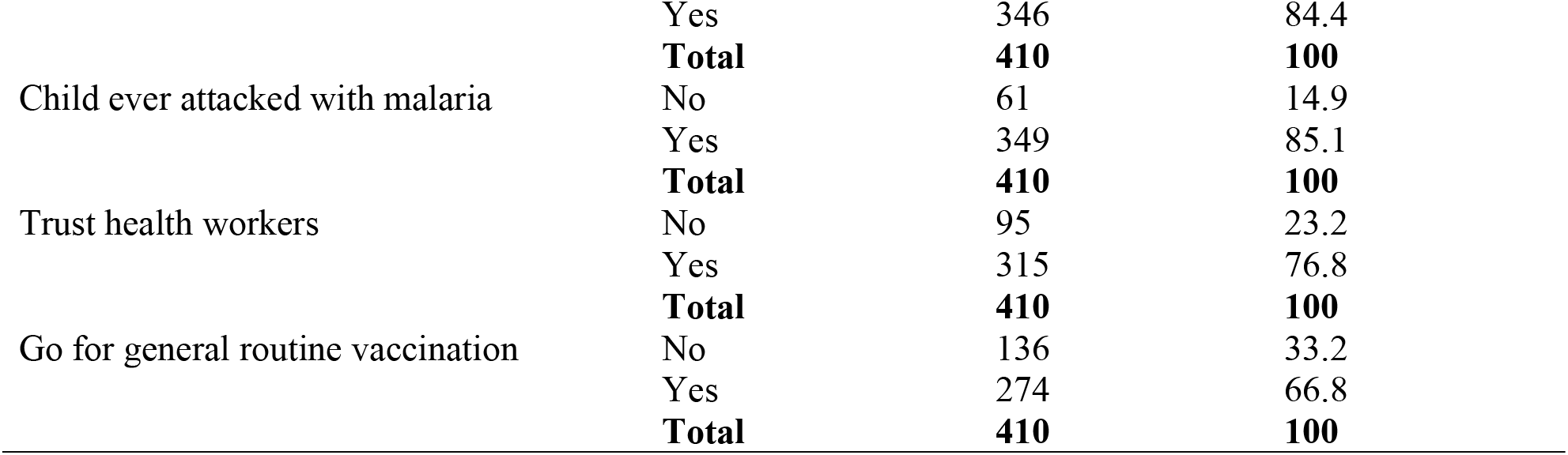
Socio-demographic characteristics of caregivers.

### Practices of under-five caregivers toward the malaria vaccine

Regarding the caregivers’ practices towards the malaria vaccine (Table 2), 278 (67.8%) children were vaccinated against. Of the 132 whose children were vaccinated, 109 (82.6%) reported keeping vaccination records and 98 (74.2) reported following up when their children missed a vaccination schedule. A total of 60 (44.8%) of those who children had taken the vaccine had taken 3 doses (Figure 1).

**Table 2:** Practices of under-five caregivers toward the malaria vaccine.

| Variable | Category | Frequency | Percentage |
| --- | --- | --- | --- |
| Child is vaccinated | No | 278 | 67.8 |
|  | Yes | 132 | 32.2 |
|  | **Total** | **410** | **100** |
| Keep vaccination records | No | 23 | 17.4 |
|  | Yes | 109 | 82.6 |
|  | **Total** | **132** | **100** |
| Follow up when child missed vaccination schedule | No | 34 | 25.8 |
|  | Yes | 98 | 74.2 |
|  | **Total** | **132** | **100** |
| Regularly seek information about malaria vaccine from reliable sources | No | 78 | 59.1 |
|  | Yes | 54 | 40.9 |
|  | **Total** | **132** | **100** |
| Ensures that child complete recommended vaccine doses | No | 37 | 28.0 |
|  | Yes | 95 | 72.0 |
|  | **Total** | **132** | **100** |
| you actively participate in community vaccination programs that offer the malaria vaccine to children | No | 85 | 64.4 |
|  | Yes | 47 | 35.6 |
|  | **Total** | **132** | **100** |
| I seek medical advice when child experiences negative symptoms after vaccination | No | 62 | 47.0 |
|  | Yes | 70 | 53.0 |
|  | **Total** | **132** | **100** |

**Figure 1:**
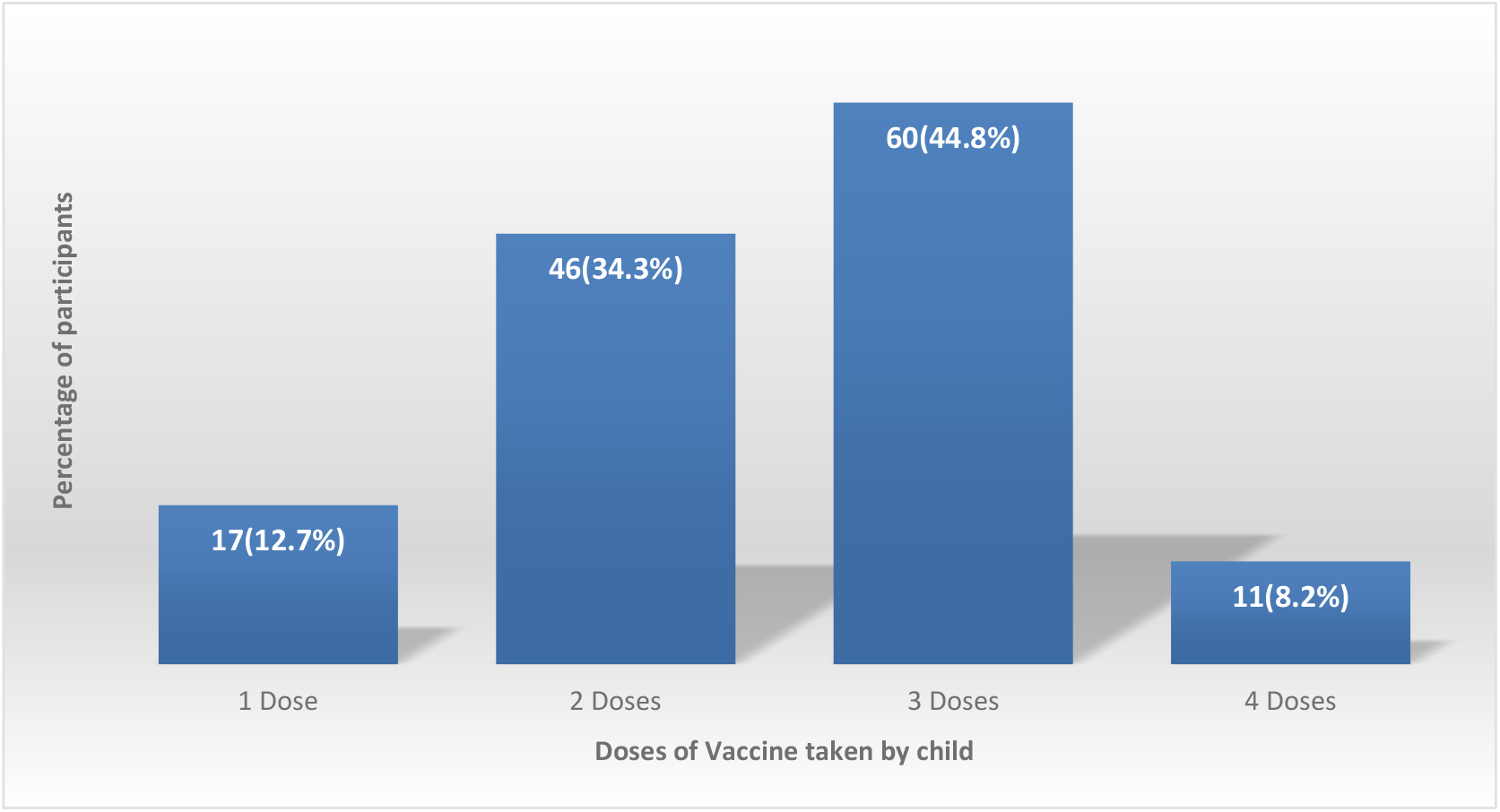
Distribution of vaccinated children by number of doses taken.

Regarding the overall practices of caregivers of under-five children towards the malaria vaccine, they were asked a total of seven questions with a maximum obtainable score of 7. The cutoff point for good practices was 60%. Hence, those who scored from 4 and above were classified as having overall good practices and those who scored below 4 as having poor practices. The overall good practices were 24.1% (Figure 2).

**Figure 2:**
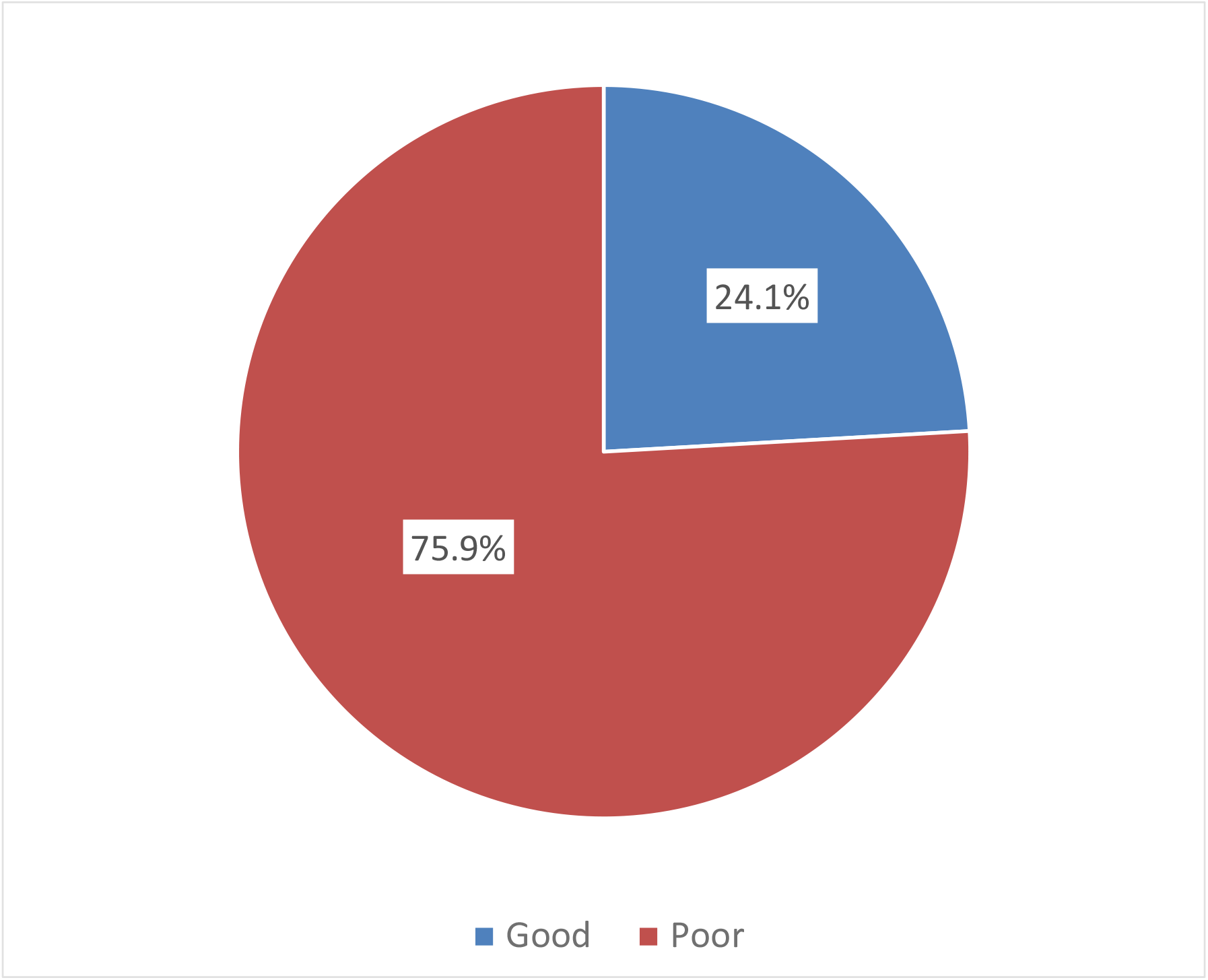
Overall practices of under-five caregivers toward the malaria vaccine in the Tiko Health District.

### Factors associated with vaccine uptake among under-five children

Regarding factors associated with the malaria vaccine uptake (Table 3), ten factors were founded significantly associated with malaria vaccine uptake in the bivariable analysis using simple logistic regression analysis. These factors were sex, marital status, occupation, relationship with child, household income, accessibility of health services, previous history of malaria, source of information, trust in health workers, and going for general child vaccination.

**Table 3:**
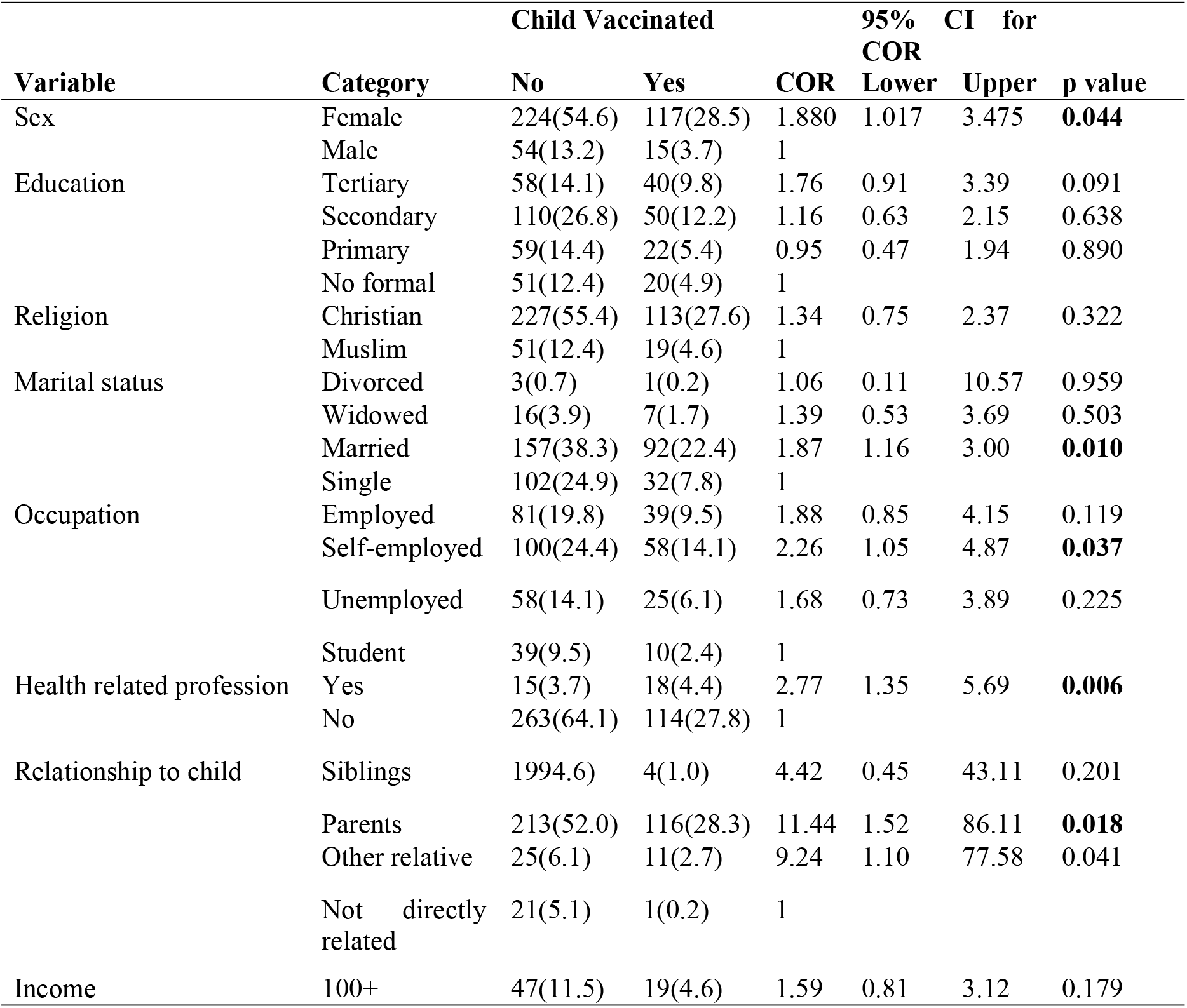
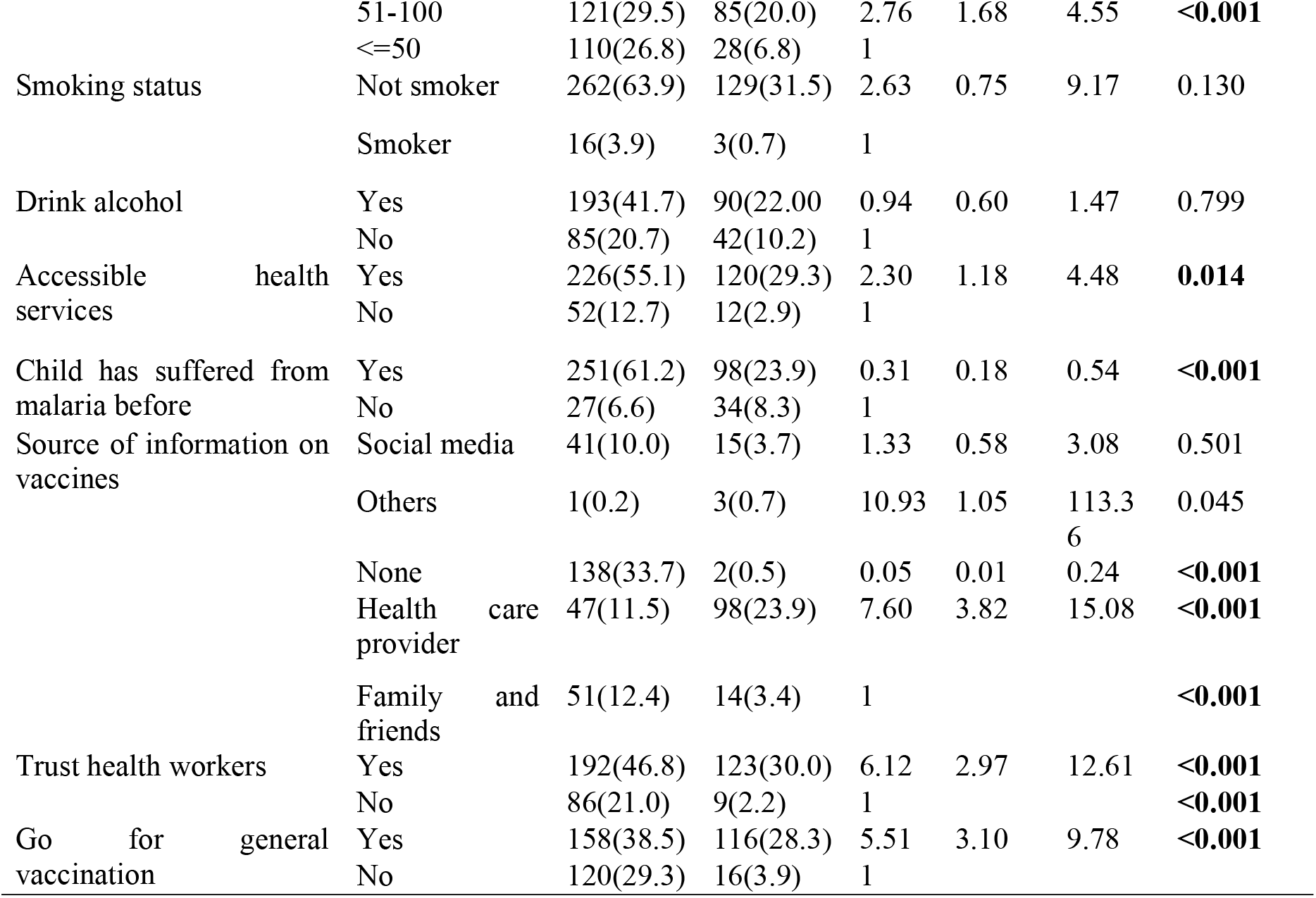
Factors associated with vaccine uptake among under-five children using simple logistic regression.

### Factors independently associated with malaria vaccine uptake among under-five children

Regarding factors independently associated with the malaria vaccine uptake by under five children (Table 4), eight factors were found independently associated with the vaccine uptake. These were; sex, profession, relationship with child, income, accessibility of health services, history of malaria infection, source of information, and trust in health workers. Under five children of female caregivers were about four times more likely to be vaccinated against malaria (aOR:4.16, 95%CI:1.47-11.75; p=0.007) as compared to children of male caregivers. Children of caregivers who were their direct parents were about eleven times more likely to be vaccinated aOR:11.44, 95%CI:1.52-86.11; p=0.008) as compared to those whose caregivers were not directly related to them. Household income was generally positively associated with vaccine uptake with children whose caregiver average monthly income was between XAF51,000 to XAF100,000 being more than two times more likely to be vaccinated (aOR:2.76, 95%CI:1.68-4.55; p<0.001) as compared to children of caregivers whose average monthly income was less than XAF50,000. Children of caregivers who reported easily accessible health services were about two times more likely to be vaccinated (aOR:2.3, 95%CI:1.18-4.48; p=0.020) as compared to those who reported not having easily accessible health services. Caregivers who reported history of malaria infection among their children were 69% less likely to have their children vaccinated (aOR:0.31, 95%CI:0.18-0.54; p<0.001). Children of caregivers who main source of information on vaccination were more than seven times more likely to be vaccinated (aOR:7.60, 95%CI: 3.86-15.08, p<0.001) as compared to those whose main source of information was family/friends. Children of caregivers who reported trusting healthcare workers were about six times more likely to be vaccinated (aOR:6.12, 95%CI:2.97-12.61) as compared to those who reported not having trust in healthcare workers.

**Table 4:**
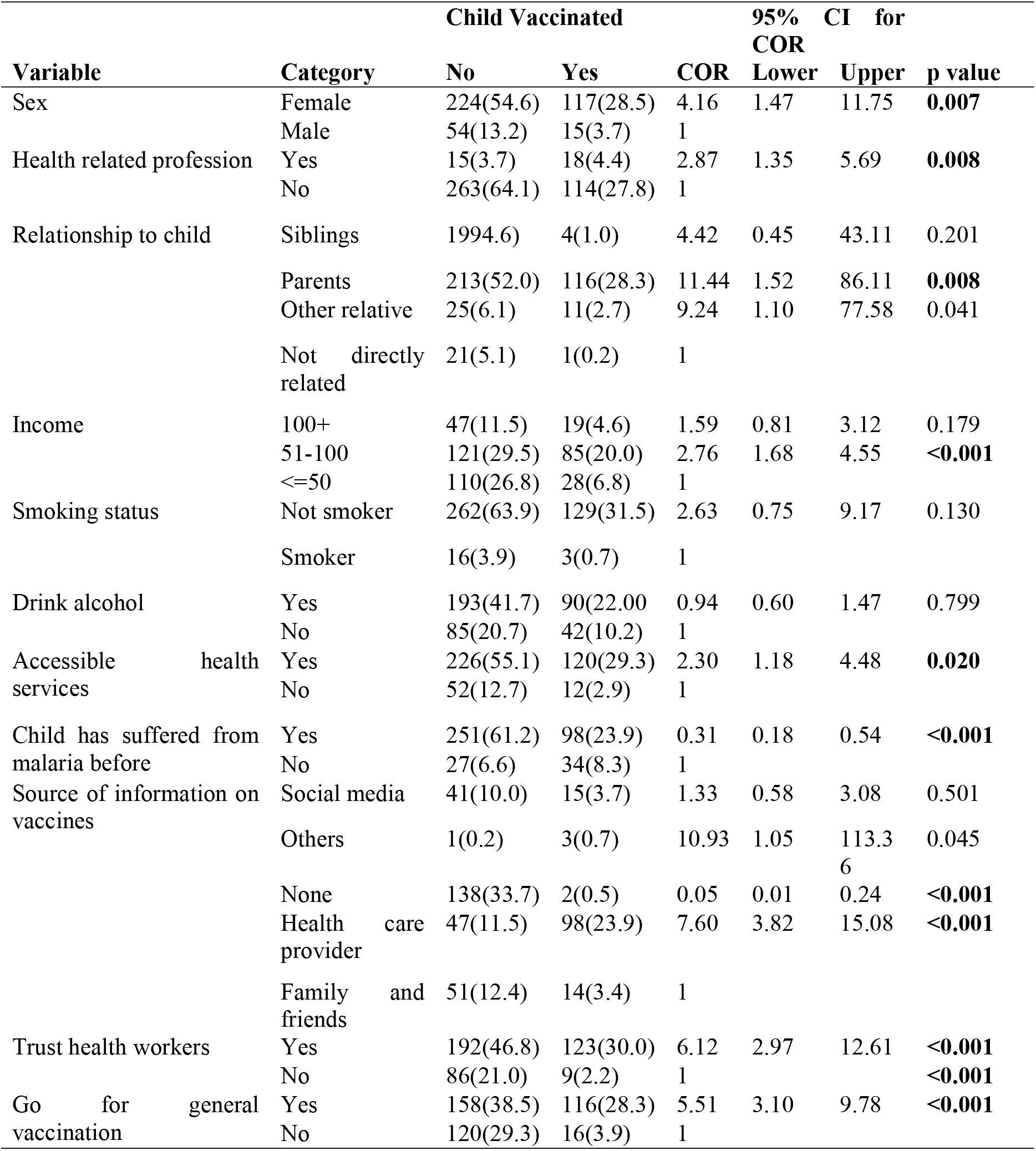
Factors independently associated with malaria vaccine uptake among under-five children.

## Discussion

The findings from this study provides important understanding in to the uptake of malaria vaccine, its associated factors and practices of under-five caregivers towards the malaria vaccine. The study found that 32.2% of caregivers had vaccinated their under-five children against malaria, this indicates a low uptake of the malaria. These findings are similar to early uptake rates in pilot countries such as Ghana, Kenya, and Malawi, conducted in 2024, where initial community response to the malaria vaccine was low [14]. Among the 132 caregivers whose children received the malaria vaccine, a high proportion 82.6% reported keeping vaccination records, which is an encouraging level of health responsibility. Similarly, 74.2% followed up when their child missed a vaccination appointment. These practices are important for ensuring full adherence to the vaccine, especially considering that RTS,S requires a multi-dose for optimal effectiveness [15]. Only 44.8% of vaccinated children had received at least three doses, suggesting challenges with vaccine completion, these results align with findings from Zambia and Tanzania, where multi-dose vaccine schedules were difficult to complete due to logistical and socioeconomic constraints [16,17]. Only 35.6% of the caregivers participated in community-based malaria vaccination programs, reflecting a concerningly low level of public engagement. Community participation is essential for achieving herd immunity and sustaining immunization coverage in high-transmission settings [18]. This study found that only 24.1% of caregivers demonstrated good overall practices regarding the malaria vaccine, indicating a generally low level of engagement and adherence to vaccination guidelines among caregivers in the Tiko Health District, the low level of good practices may be attributed to several factors, including limited health education, poor awareness of vaccine schedules, and low community-level sensitization at the time of vaccine rollout. This study identified several variables that were independently associated with malaria vaccine uptake among children aged 0–5 years in the Tiko Health District, which were; sex, profession, relationship with child, income, accessibility of health services, history of malaria infection, source of information, and trust in health workers. Children whose primary caregivers were female were four times more likely to be vaccinated compared to those cared for by males. This confirms findings from previous studies indicating that mothers are typically more involved in child health decisions and are more likely to engage in preventive healthcare measures such as vaccination [19]. In many African settings, traditional gender roles place child health responsibilities largely on women, who often accompany children to health facilities [20]. Caregivers working in health-related professions were three times more likely to vaccinate their children. This is could due to the fact that, those with medical or health training are more likely to understand the benefits of vaccines, perceive their safety positively, and access services more readily [21]. This aligns with WHO’s assertion that health literacy among caregivers significantly improves vaccine confidence and completion [1]. Children living with their biological parents were significantly more likely to be vaccinated than those under the care of other relatives or unrelated guardians. This finding is supported by research in Tanzania and Kenya showing that non-parental caregivers often have lower prioritization of child health, less consistent healthcare-seeking behavior, and reduced vaccination rates [22]. Higher household income was positively associated with vaccine uptake. Children from households with a monthly income between XAF 51,000 and XAF 100,000 were more than twice more likely to be vaccinated compared to those from households earning less than XAF 50,000. This is similar to a study carried out in Cameroon, where economic stability was found to facilitates better access to healthcare services and adherence to vaccination schedules [23]. Caregivers who reported easy access to health services were about twice more likely to have their children vaccinated. Accessibility plays a crucial role in healthcare utilization, and efforts to improve healthcare infrastructure and reduce travel barriers are essential for increasing vaccine coverage [24]. Caregivers who received information about the malaria vaccine from healthcare providers were more likely to vaccinate their children. Trusted sources of information, such as healthcare workers, play a vital role in influencing health behaviors. In Cameroon, community-based risk communication involving health workers has been effective in promoting vaccine acceptance [24]. Trust in healthcare workers was a strong predictor of vaccine uptake. Caregivers who trusted health workers were about six times more likely to vaccinate their children, building and maintaining trust through consistent, transparent, and culturally sensitive communication is essential for the success of vaccination programs [24].

### Recommendations

To improve caregiver practices toward malaria vaccination, targeted community sensitization and health education campaigns should be implemented to increase awareness of vaccine schedules, benefits, and the importance of completing all required doses. Efforts should focus on both female and male caregivers to promote shared responsibility in child healthcare.

To address the determinants associated with malaria vaccine uptake, strategies should be put in place to improve access to vaccination services, particularly for low-income households and those in hard-to-reach areas. Strengthening trust in healthcare workers through community engagement and training in effective communication is also essential.

### Conclusion

The uptake of malaria vaccine in the Tiko Health District was low with only 32.2% of children having received at least one dose of the malaria vaccine. The overall good practice was 24.1%, which was low. Several factors were found to be independently associated with vaccine uptake, including caregiver sex, profession, relationship with the child, household income, accessibility of health services, history of malaria infection, trust in healthcare workers, and source of information.

## Data Availability

Data Availability Statement: The data that support the findings of this study are available from the corresponding author upon reasonable request. The data have been anonymized to maintain participant confidentiality and are stored in a secure, password-protected repository.

## Authors Contribution

**Conceptualization**: Idang Maureen Abiache, Divine Nsobinenyui, Chrisantus Eweh Ukah, Yunika Larissa Kumenyuy, Ngu Claudia Ngeha, Randolf Wefuan, Syveline Zuh Dang, Ndip Esther Ndip, Mirabelle Pandong Feguem, Dickson S. Nsagha

**Data Curation**: Idang Maureen Abiache, Divine Nsobinenyui, Chrisantus Eweh Ukah, Yunika Larissa Kumenyuy, Ngu Claudia Ngeha, Randolf Wefuan, Syveline Zuh Dang, Ndip Esther Ndip, Mirabelle Pandong Feguem, Dickson S. Nsagha

**Formal Analysis:** Idang Maureen Abiache, Divine Nsobinenyui, Chrisantus Eweh Ukah, Yunika Larissa Kumenyuy, Ngu Claudia Ngeha, Randolf Wefuan

**Investigation**: Chrisantus Eweh Ukah, Nicholas Tendongfor, Alan Hubbard, Elvis A. Tanue, Rasheedat Oke, Nahyeni Bassah, Sandra I. McCoy, Larissa Kumenyuy Yunika, Claudia Ngeha Ngu S. Ariane Christie, Dickson S. Nsagha, Alain Chichom-Mefire, Catherine Juillard

**Methodology:** Idang Maureen Abiache, Divine Nsobinenyui, Chrisantus Eweh Ukah, Yunika Larissa Kumenyuy, Ngu Claudia Ngeha, Randolf Wefuan, Syveline Zuh Dang, Ndip Esther Ndip, Mirabelle Pandong Feguem, Dickson S. Nsagha

**Supervision:** Abiache, Divine Nsobinenyui, Chrisantus Eweh Ukah,

**Validation:** Divine Nsobinenyui, Chrisantus Eweh Ukah, Dickson Shey Nsagha

**Visualization**: Chrisantus Eweh Ukah, Yunika Larissa Kumenyuy, Ngu Claudia Ngeha, Randolf Wefuan, Syveline Zuh Dang, Ndip Esther Ndip

**Writing – original draft:** Idang Maureen Abiache, Divine Nsobinenyui, Chrisantus Eweh Ukah, Yunika Larissa Kumenyuy, Ngu Claudia Ngeha, Randolf Wefuan, Syveline Zuh Dang, Ndip Esther Ndip, Mirabelle Pandong Feguem

**Writing – Review and Editing:** Idang Maureen Abiache, Divine Nsobinenyui, Chrisantus Eweh Ukah, Yunika Larissa Kumenyuy, Ngu Claudia Ngeha, Randolf Wefuan, Syveline Zuh Dang, Ndip Esther Ndip, Mirabelle Pandong Feguem, Dickson S. Nsagha

